# Naturalistic sleep tracking in a longitudinal cohort: how long is long enough?

**DOI:** 10.1101/2024.10.19.24315818

**Authors:** Balaji Goparaju, Glen de Palma, Matt T. Bianchi

## Abstract

**Background:** Despite broad interest in the health implications of sleep duration, traditional measurements via polysomnography or actigraphy are often limited to one or a few nights per person. Given the potential variability of sleep duration over time, inferential uncertainty remains an important issue for relatively short observation windows.

**Methods:** We describe potential limitations of shorter duration sleep tracking by sub-sampling from longer-term observation windows, using a combined approach of simulated data from known distributions, in addition to real-world data (30-365 nights) from over 35,000 participants who provided informed consent to participate in the Apple Heart and Movement Study and elected to contribute sleep data to the study.

**Results:** Simulations demonstrate that the magnitude of deviation from truth, defined using all available observations per individual, as well as the presence and direction of bias, depended on the sub-sample size, the type of simulated distribution (Gaussian versus skewed), and the summary statistics of interest, such as central tendency (mean, median) and dispersion (standard deviation (SD), interquartile range). For example, the SD computed from n=7 observations from a simulated normal distribution (7±1 hours) showed a median 6.7% under-estimation bias (IQR 24% under- to 14.7% over-estimation). Real-world sleep duration data, when under-sampled and compared to longer observations within-participant, showed similar SD bias at 7 nights, and similar convergence rates approaching the true value (based on 90 nights) as longitdunal sample number increases. Shapiro-Wilk tests for normality and log-normality show that 64% of simulated log-normal (skew) distributions fail to reject normality at n=7 samples, while real-world sleep duration data most commonly failed both normality and log-normality tests. Finally, simulated cohorts with sleep durations of 7±1 hours mixed with a subset of 6±1 hours sleepers showed that a random single-night observation of “short sleep” (6 hours) is more likely from random variation of a 7-hour sleeper, than from an actual 6-hour sleeper. Extending the observation to n=7 nights mitigates this mis-classification risk.

**Conclusion:** The results of simulations and empiric data patterns suggests that longer duration tracking provides important and tangible benefits to reduce bias and uncertainty in sleep health research that historically relies on small observation windows.

## 1. Introduction

Although sleep is widely understood as a pillar of health and wellbeing, the objective measurement of sleep in research studies has been rather limited to short duration tracking via either gold-standard laboratory polysomnography (PSG) or approximations of sleep via actigraphy in the real-world. For example, the epidemiology of sleep stage architecture, described in two meta-analyses, is derived mainly from 1-3 nights of PSG data^1,2^. Similarly, in a recent review focused on nightly variability of sleep, actigraphy studies were mostly 7-14 nights in duration, with only seven of the 53 studies using actigraphy for more than 14 nights (and of those, the largest sample size was 141 subjects)^3^. Given the nightly variability in sleep duration, in part due to multiple biological, sociological, and behavioral influences on sleep^4^, capturing 1-14 nights may be an under-sampled view of a person’s sleep patterns.

Even the use of self-reported diary entries, which in principle could continue for extended periods, are often used for relatively short windows into sleep patterns. For example, the insomnia clinical guideline recommends a “two-week sleep log to identify sleep-wake times, general patterns, and day-to-day variability”^5^. The one-week consensus sleep diary, which was designed primarily for insomnia research but is general in its content, does not indicate how many weeks ought to be tracked^6^. The health and wellbeing implications of both self-reported and objective sleep tracking may be limited by short duration observation windows, with little data-driven evidence to rationalize any particular duration. Quantifying some of the statistical uncertainties that might accompany relatively short duration tracking could improve a range of research efforts in this space.

To frame the fundamental question, “how long is long enough?”, we used a combination of two approaches: 1) simulations with known distributions, and 2) real-world objective longitudinal data from participants who consented to participate in the Apple Heart and Movement Study^7,8^. We hypothesized that sub-sampling a range of nights from individuals with long-term objective sleep observations would help quantify uncertainty and bias when measures of centrality and dispersion are computed from short duration samples. Sub-sampling from simulated distributions provides bounding context under simplifying assumptions, i.e., uncertainty and/or bias attributable solely to sub-sampling. These results set the stage to then interpret sub-sampling results from empiric sleep data, where neither the true distribution nor its stationarity can be assumed. For comparison with a different longitudinal health tracking metric, we also performed sub-sampling of objective exercise data from the same study cohort. The results provide statistical inference context when interpreting small observation windows of any health metric that varies over time. The simulated and empiric data analysis suggests that the question of “long enough” does not have a singular or simplistic answer. Instead, it depends upon multiple factors, such as the underlying data distribution, the summary metric of interest (e.g., measures of central tendency vs measures of dispersion). From a pragmatic perspective, sub-sampling across a range of duration provides estimates of convergence rates, such that hypothesis-specific goals can be considered in terms of trade-offs of participant burden and how close to the theoretical truth a given observation window might be.

### 2. Methods

#### 2.1 AHMS overview

This study included participants who consented to participate in the Apple Heart and Movement Study (AHMS)^8^, and opt in to sharing HealthKit data streams including sleep tracking data. This population includes adults residing in the United States who own an iPhone and an Apple Watch. This study collects a variety of data types including objective sleep and exercise data via HealthKit. Participants opt in to sharing data, and can withdraw from the study at any time.

#### 2.2 Simulations

Simulations were performed in Microsoft Excel and plotted with GraphPad Prism. Normal distribution data was generated in Excel, using the function NORM.INV(RAND(),mean,SD), and skewed distributions were generated in Excel using the functions BETA.INV(RAND(), alpha, beta) or LOGNORM.INV(RAND(), mean, SD). For example, in the normal distribution simulation for TST, a mean of 7 hours and standard deviation (SD) of 1 hour would be similar to self-reported variability in adults^9^ and similar to that seen in the AHMS cohort.

#### 2.3 Study population

AHMS was approved by the Advarra Central Institutional Review Board, and registered to ClinicalTrials.gov (ClinicalTrials.gov Identifier: NCT04198194). Informed consent is provided within the Apple Research app. Participation does not require sleep tracking. However a subset of participants do track and opt in to sharing sleep data. In the current work, we restricted sleep analysis to when the data source was the first-party sleep application on the Apple Watch. To be included in the current analysis, we pre-defined the minimum data requirements and time window. For the sleep and exercise analysis, we required at least 90 days with sleep duration of at least 4 hours, in a window between Sept 1, 2021, and September 1, 2022 (n=40,441). Sleep duration values <4 or >13 were excluded, and exercise values <3 or >300 were excluded. For the sleep stages analysis, we required at least 30 days with sleep duration of at least 4 hours in a window between October 15, 2022, and April 15, 2023 (n=36,956 participants), to correspond to the availability of first party Watch sleep staging. In both cases, only sleep data from first party Apple Watch source was included.

#### 2.4 Data Types

Sleep data was obtained from HealthKit, and the total sleep duration was computed for each night, using only Apple Watch first party sleep tracking data. During the time period analyzed for the sleep and exercise data (2021 to 2022), Apple Watch sleep tracking provided a binary output, asleep or not, for those onboarded to the sleep experience and wearing their watch to track sleep. As of iOS 16 and WatchOS 9, the sleep feature was updated to include sleep stage information, for a total of four states (awake, Core Sleep, Deep Sleep, REM sleep)^10^.

Exercise data was obtained from HealthKit, as the number of minutes of exercise, which is captured in one of the three activity “rings” on the Apple Watch. Capturing exercise minutes happens passively during routine watch wear. User-indicated workouts entered using the Workout app also contribute to exercise minutes, but we made no distinction in the current analysis whether exercise minutes came from a workout or not; we simply assess the total minutes in HealthKit per day. This daily value is the sum of exercise minutes from midnight to midnight.

#### 2.5 Analysis of real-world health data

Statistical analyses and plotting were done in Python for sleep and exercise data from AHMS participants. Where p-values are incorporated (for tests of normality and log-normality), we used a 0.05 threshold for significance. When sub-sampling was performed in simulated data, the values are independent by definition, such that sub-sampled points can be considered part of a stationary process. By contrast, empiric sleep and exercise data from AHMS participants cannot be assumed independent or stationary. Thus, for the empiric data sub-sampling, random days were chosen as opposed to sequential days. As such, certain non-stationarities would not be captured such as weekend effects or other transient changes (e.g., illness, vacation).

### 3. Results

#### 3.1. Simulations

To provide context for interpreting basic descriptive statistics based on a limited number of observations, we framed the approach as one of comparing sub-sampled observations to an individual-level “ground truth” computed from longer duration observations from the same individual. We began with simulated distributions under two simplifying assumptions: known statistical distributions, and independence of observations within each simulated individual (no temporal relationships, i.e., the time series is stationary). We performed simulations with four pre-defined distributions: two normal distributions (same mean, but different SD values), and two skewed beta distributions with opposite skew directions (**Figure 1A**). Using these data generating functions, we simulated 5,000 individuals for the four distributions, with each individual having 1,000 values (“days”) of data with arbitrary units, from which a series of descriptive statistics were computed by sub-sampling from each simulated individual. The ground-truth values were defined by the result of using all 1,000 values per simulated individual. The results of sub-samples for n=7, n=14, and n=30 nights are given for the mean, median, SD, and IQR calculations (**Figure 1B-E**). Additional sub-sampling from Gaussian and log-lormal simulations, ranging in duration from 3-720 days, are shown in **Supplemental Figure S1**.

**Figure 1.**
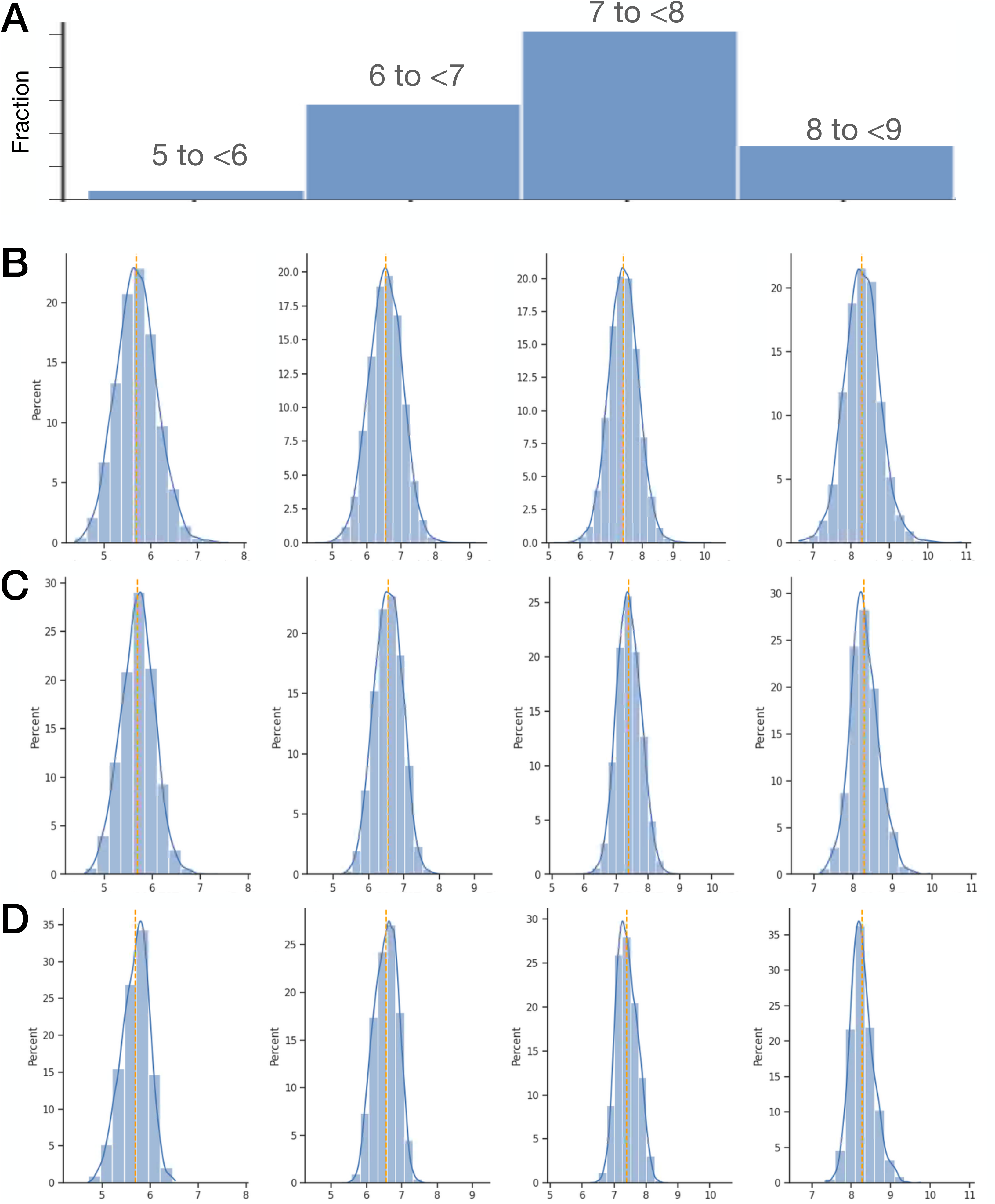
Descriptive statistics computed from sub-samples drawn from two normal and two skewed simulated distributions. A. Frequency histograms from four simulated distributions: two Gaussian distributions with same mean (100 a.u.) but different SD values (15 vs 30 a.u.), and two skewed beta distributions with right vs left tails (right tail with alpha 0.5, beta 3.0, and left tail with alpha 3.0 and beta 0.5), scaled to be in the range of 0-100 a.u. The plots are derived from 5000 samples in each case, with bin size of 10 or 5 for normal vs skewed distributions, respectively. The next four rows show computed measures of centrality (mean (B) and median C)) and dispersion (SD (D) and IQR (E)), computed from sub-samples drawn from the corresponding distributions shown in row A. In each panel, random draws of size n=7, n=14, or n=30, are summarized as the mean (circles) and standard deviation (error bars; SD) computed from of n=5000 simulated subjects. The summary metrics are normalized and reported as a percentage of the value obtained from n=1000 draws for each subject, taken to be the reference truth (dotted line at Y=100%).

Several observations are notable from these simulations. For the two normal distributions, uncertainty in the mean and median are clearly proportional to the SD (column 1 versus column 2), and no systematic directional bias is evident. However, uncertainty in measures of dispersion (SD and IQR) were independent of the SD for the two normal distributions. Furthermore, the SD and IQR both exhibit under-estimation bias for the smaller sub-samples, which was more prominent in the IQR calculations. Additional sub-sampling simulations and associated measures of dispersion from sub-samples of Gaussian distributions with different SD values are given in **Supplemental Figure S2**.

For the skewed distributions, the mean and median exhibited distinct behavior depending on the direction of the tail. In the rightward tail simulations, the median contained markedly increased uncertainty (error bar span over twice as large as for the mean), and also showed a directional bias toward over-estimation (whereas the mean showed no such bias). The SD and IQR both show an under-estimation bias, slightly more prominent in the IQR than in the SD. In the leftward tail simulations, the SD and IQR showed similar behaviors as compared to the rightward tail (more prominent under-estimation in IQR than in the SD). The mean and median showed much smaller ranges of uncertainty. This is attributed to the relative nature of the %-of-truth computation, where the same absolute difference from truth in a left versus right tailed skew corresponds to a distinct percent of truth.

Comparing the Gaussian distributions to the right skewed distributions, the relative uncertainty (error bars across the 5,000 simulations) was larger for the right skewed distribution regarding measures of centrality (**Figure 1B**, **1C**). By contrast, uncertainty in the measures of dispersion were only modestly larger in the skewed compared to the Gaussian simulations (**Figure 1D, 1E**). For additional context, different variations on measures of dispersion are shown for a normal distribution in **Supplemental Figure S3**.

#### 3.2. Apple Heart and Movement Study: Longitudinal sleep duration and exercise data

The above sub-sampling simulations highlight uncertainties in basic descriptive metrics attributed solely to sub-sampling, since the distribution shapes are known and stationary. In real-world settings, neither the true distribution nor stationarity can be assumed. When longitudinal data is available, we have a unique opportunity to employ a similar empiric approach via within-participant under-sampling, to answer questions like, “if we only had 7 nights of data from each participant, how different might measures of central tendency and dispersion be, compared to using much longer observation windows?” The AHMS cohort provides such an opportunity.

**Figure 2** shows the results of random sub-sampling each individual’s sleep duration (total sleep time; TST) and exercise duration (minutes per day) values to compute descriptive statistics, where ground truth per participant is computed from at least 90 values for each metric. The TST sub-sampling shows no directional bias for mean or median computations (**Figure 2**, **A1 and A2**). However, the exercise sub-sampling shows over-estimation for the median (**Figure 2**, **B1 and B2**), reminiscent of the simulations of a right-skewed distribution. The SD and IQR both exhibited under-estimation bias at lower sub-sample values, for both sleep and exercise, before gradual convergence to within 5% of per-participant truth (**Figure 2**, **A3, A4, B3, B4**). In summary, the empiric data sub-sampling quantified directional bias and convergence pace in real-world data, especially for the windows of observation of ≤14 nights that are typical of objective sleep tracking studies.

**Figure 2.**
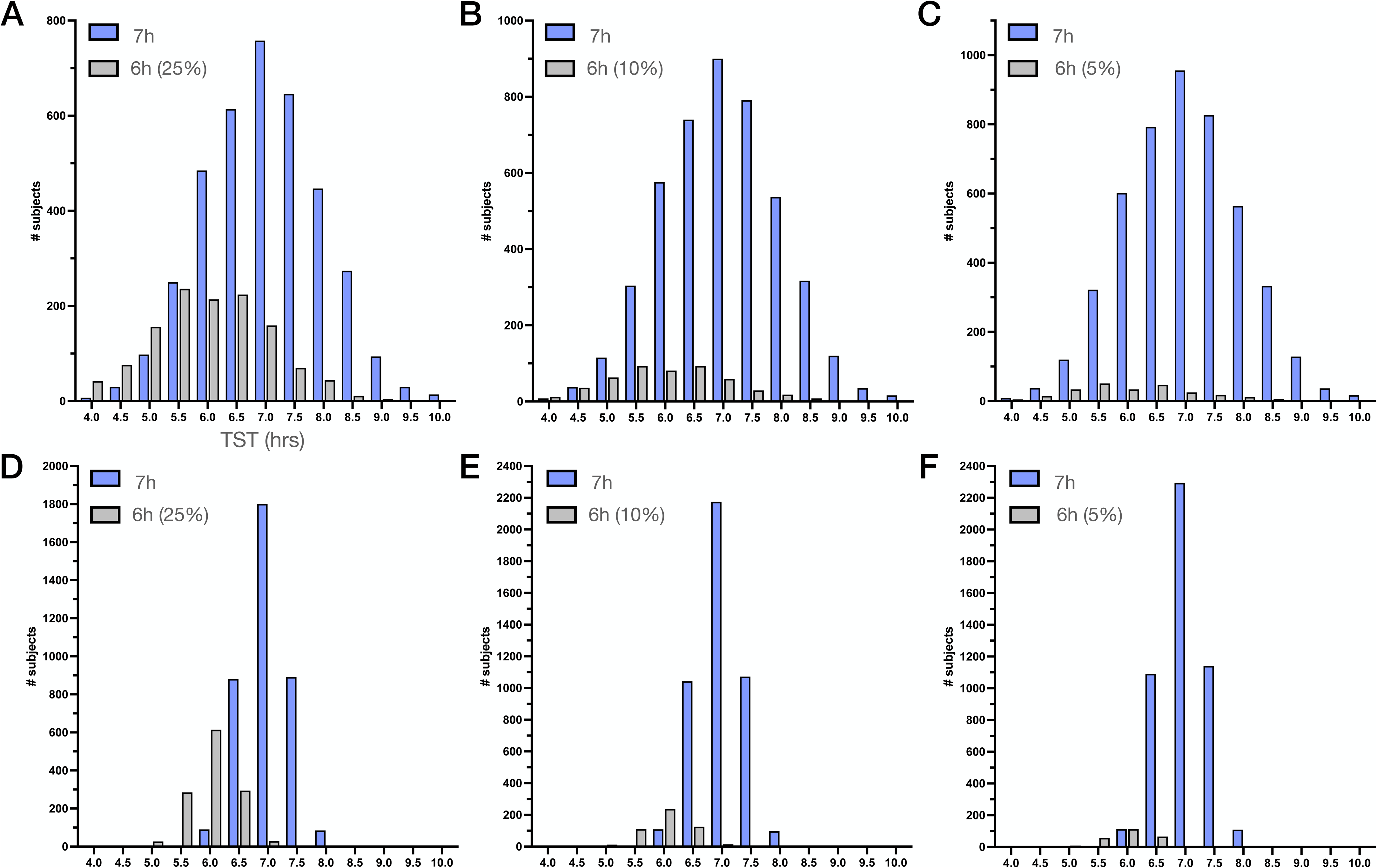
Descriptive metrics calculated across varying sample sizes of real world sleep and exercise data. In each panel, the mean (circles) and standard deviation (error bars; SD) of each summary metric is given for n=40,441 participants, where the X axis indicates the number of observations drawn from each participant (X label in A1 applies to all panels). The ground truth descriptive statistics for TST (hrs) and exercise (minutes) for each participant are determined from at least 90 values. Note the sample size on the X axis has non-linear increments, indicated by the vertical dotted lines.

#### 3.3. Apple Heart and Movement Study: Longitudinal sleep stages data

Although sleep duration and its variability are common metrics used in epidemiology studies, some study designs use PSG testing to allow for stages within sleep to be assessed as another window into sleep physiology. PSG testing in large cohorts is even more limited in duration than actigraphy, with typically 1-3 nights of PSG assessment per participant, as in the two largest meta-analyses of sleep staging data from PSG^1,2^. We sought to answer a similar question for stages as we did for duration: “how different might an estimate of sleep stage duration (or %) be, from sub-samples of size 1, 7, 14, and 30 nights, as compared to estimates from longer durations of data?” **Figure 3** shows this analysis from empiric sleep data, for stage duration and percentage. For each participant, the values computed from sub-samples are compared relative to a within-participant ground truth computed from longer observation windows. A sub-sample size of n=1 night showed the most divergence, as expected, with median absolute difference of approximately 20 minutes for REM (and about 25% showing 30+ minutes of deviation). The median absolute divergence was approximately 10 minutes for Deep and Wake stages. The median divergence of REM as a percentage of TST was ∼3.5%. For context, this change in REM sleep is larger than the 3% difference in REM percentage reported in a cohort study associating REM percentage to dementia using a single night of PSG^11^. The observed divergence from “ground truth” reduces proportionately as the computations are made using 7, 14, or 30 nights to compute stage information within-participant.

**Figure 3.**
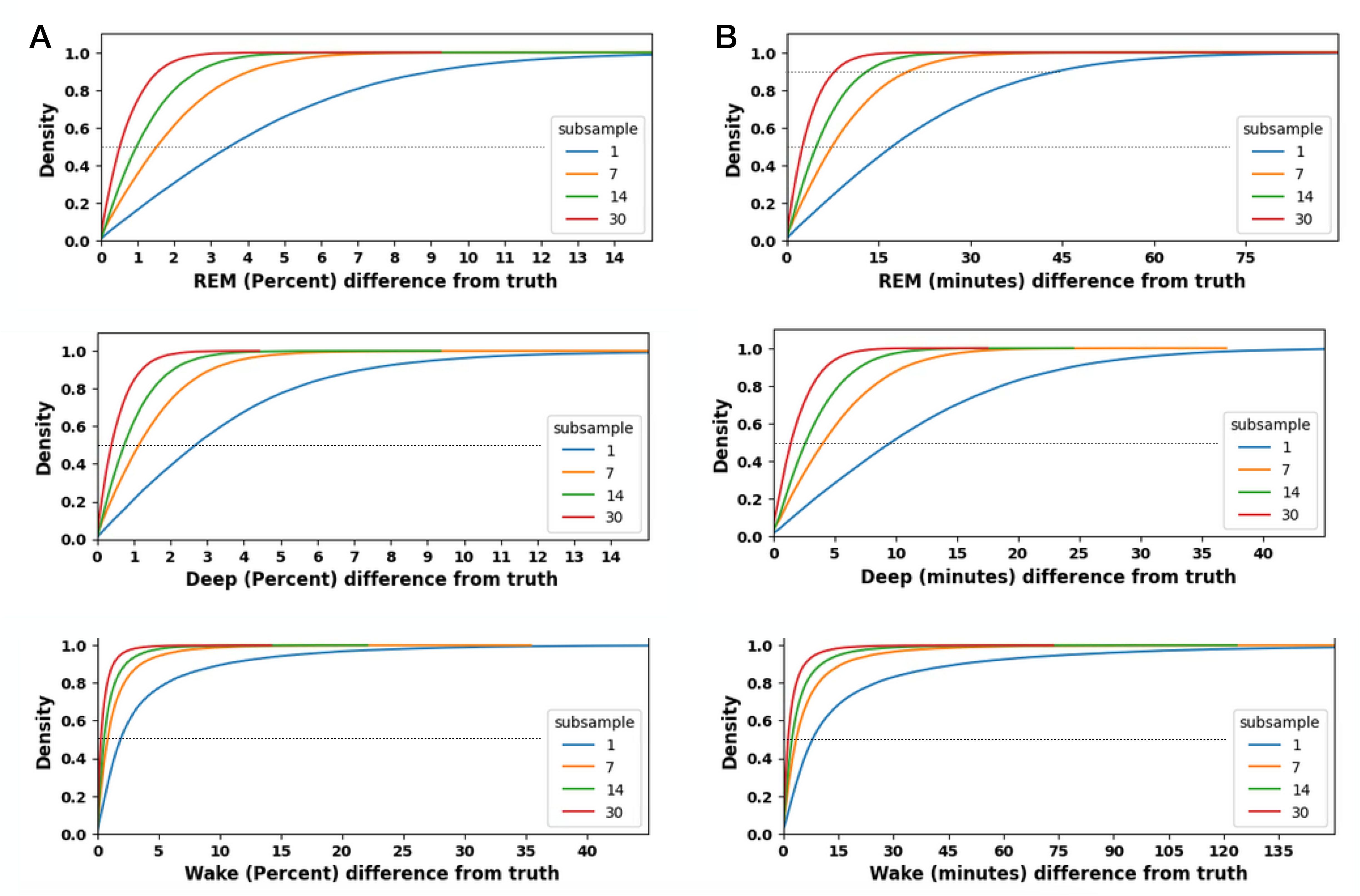
REM, Deep, and Wake amounts computed from sub-samples drawn from real-world sleep tracking. In each panel, CDF curves are plotted for sub-samples of size 1 (blue), 7 (orange), 14 (green), or 30 (red) nights, compared to the true amount within participant computed from at least 90 nights. The X axis indicates the percent (A) or the absolute amount (B) of REM sleep (first row), Deep sleep (second row), or wake after sleep onset (third row). The horizontal dotted line in each panel corresponds to the 50th percentile (density of 0.5); the intersection of each CDF with that line can be interpreted as half of the observations have at least that X-value of stage deviation from truth, or more.

#### 3.4. Assessing normality and log-normality in simulated and real-world data

In any setting of limited observations, the distribution shape may be challenging to ascertain. As a result, statistical decisions such as whether to the use a mean versus a median to summarize central tendency may be unclear. To frame this challenge, we first simulated normal and log-normal (skewed) distributions (same as those shown in **Supplemental Figure S1**). We then applied Shapiro-Wilk tests of normality and of log-normality to these simulated individuals (**Figure 4A**). At a sub-sample of 7 or 14 days, the data sampled from a normal distribution is similarly likely to pass either test. Even at 180 days, 43.8% of the simulations from the normal distribution still pass the log-normality test. True log-normal simulations are less likely to pass normality, even at 7 nights (although about two thirds still do pass at this small sub-sample), and this normality pass-rate rapidly approaches zero by 30 days. Shapiro-Wilks tests were subsequently applied to the real-world observations of sleep duration and exercise minutes in AHMS participants. Each metric in each participant was tested for normality or log-normality with the two Shapiro-Wilks tests, using only the first 180 nights available to keep the sample size fixed for each participant (**Figure 4B**). For both TST and exercise, the most common result was to reject both normality and log-normality, with this dual-rejection result being much more common in the exercise data (>90%); exercise passed log-normality in around 10% of participants. By contrast, the % passing only normality was 10x higher for TST than for exercise, while a considerable subset of TST passed both tests. These results suggest the heterogeneity of real-world sleep and exercise distributions exceed that predicted solely by sample size from the above simulations.

**Figure 4.**
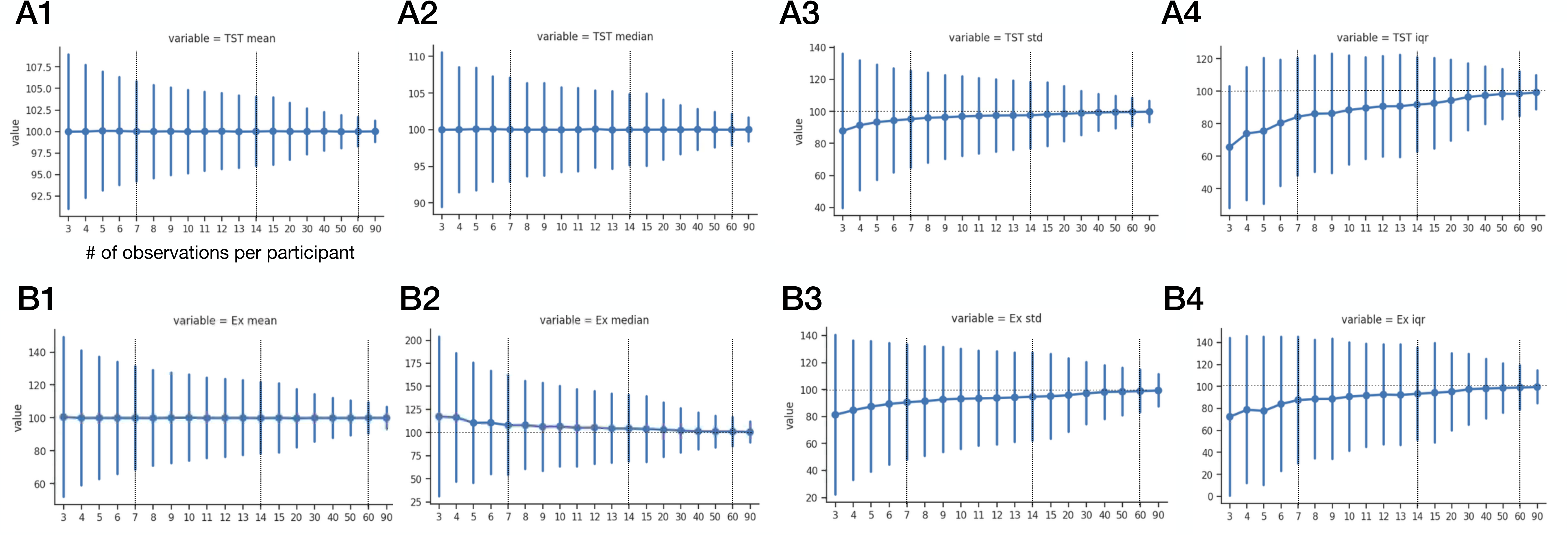
Tests of normality and log-normality, in simulated and empiric data. A. The percentage of a simulated cohort (n=5000) passing Shapiro-Wilks tests of normality and log-normality for sub-samples of 7, 14, 30, 60, and 180 values. The 4 curves are the possible combinations of normal and log-normal distributions each being tested for normality and log-normality. The legend letters indicate the source distribution first, then the type of test applied. N: normal, LN: log-normal. B. The percentage of empiric data for sleep (left bars) and exercise (right bars) that passed Shapiro-Wilks tests of normality and/or log-normality. Exactly 180 values are included per participant in this analysis.

#### 3.5. Short-sleep duration subsets: simulations and real-world data

A single value for sleep duration may sometimes be used in epidemiology settings. One such circumstance involves quantifying “short sleep” as has been done with PSG testing in the context of subjective versus objective insomnia phenotypes ^12^. In this setting of a single objective TST value, duration-based labels such as “short sleep” (e.g., less than 6 hours), may be viewed as an extreme form of under-sampling. Again, we approach this question first with simulations, and then with real-world objective sleep tracking data from AHMS participants. For the simulations, we created a mixture of two sub-cohorts, one drawn from a normal distribution with mean and SD of 7+/-1 hours, and a “short sleep” cohort drawn from a mean and SD of 6+/-1 hours. **Figure 5** illustrates the distribution histograms of TST values for each sub-cohort, across three prevalence values for the 6-hour group: 25%, 10% or 5% of the total cohort (the remaining fraction being drawn from the 7+/-1 hour distribution; **Figure 5A-C**). We can visualize and estimate the relative likelihood of an individual belonging to the 6-hour or 7-hour cohort if we assign their bin using only a single TST value. For example, consider the bin centered at 6 hours of TST: when the total cohort mixture includes 25% of the true 6-hour individuals, an observation of 6 hours is about twice as likely to come from the true 7-hour sub-cohort than from the true 6-hour sub-cohort. When the 6-hour sub-cohort represents a smaller subset, say, 10% or 5% of the total, a single observation in the 6-hour bin is 7x or 20x, respectively, more likely to have come from random variation of a true 7-hour sub-cohort individual. In other words, under these simplified but plausible assumptions, the vast majority of single-night 6-hour TST bin observations actually came from random variation of the 7-hour group, and thus could be inadvertently mis-classified as “short”. Likewise, when individuals with TST values of 6 hours or fewer are considered “short”, such observations are more likely to arise from random deviations of the 7-hour group, for all three relative prevalence simulation scenarios.

**Figure 5.**
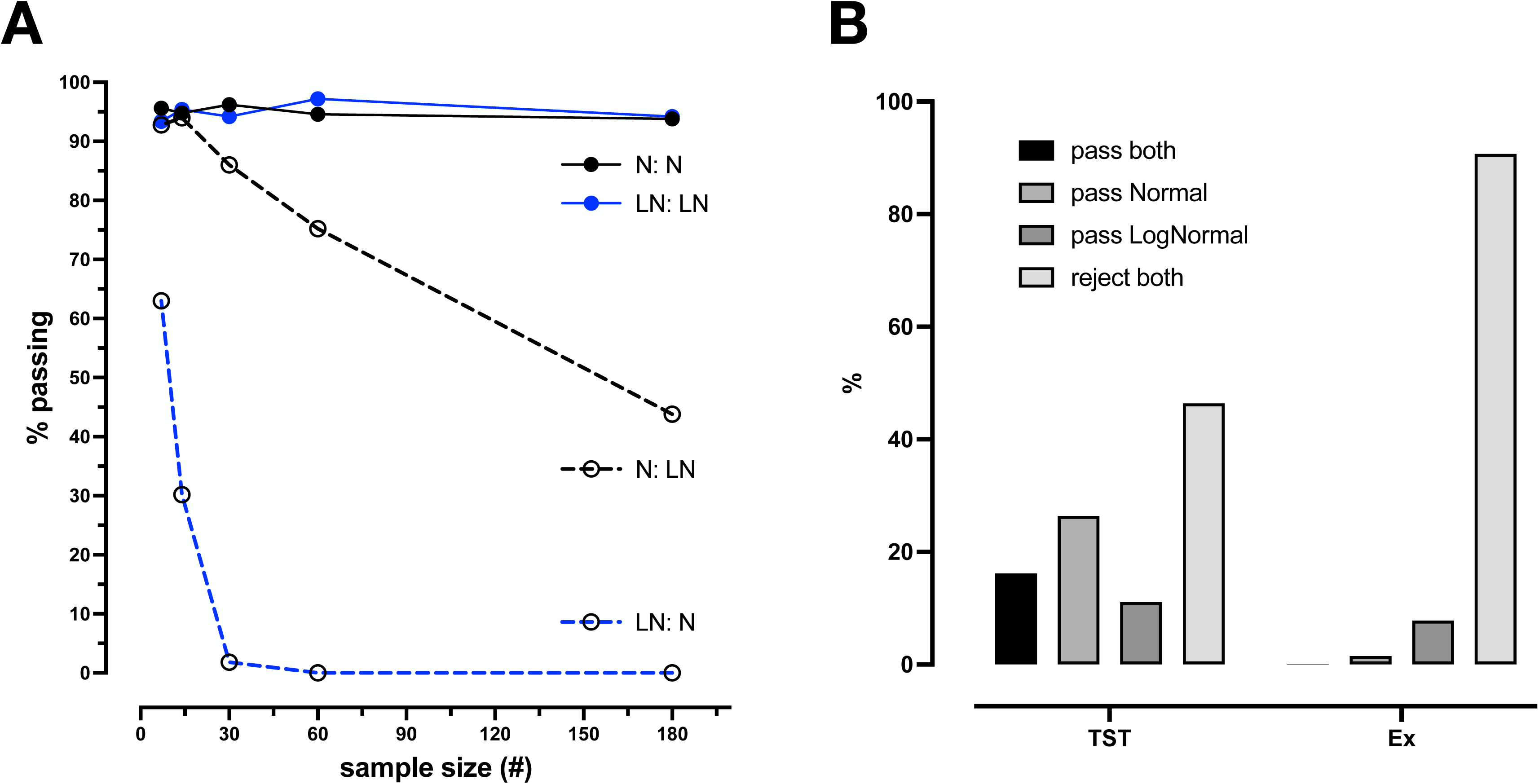
Relative distributions of TST values from a mixture of two simulated normal distributions. In each panel, the two distribution histograms are shown, in 30 minute bin widths, one for a simulated cohort with TST drawn from a normal distribution of mean and SD of 7+/-1 hours (blue), and one for a simulated cohort drawn from a normal distribution of mean and SD of 6+/-1 hours (gray). The X axis label in panel A applies to all panels. In the top row, the percentage of the total cohort (5,000) drawn from the 6-hour group is 25% (A; 1,250 subjects), 10% (B; 500 subjects), or 5% (C; 250 subjects. Each subject contributes a single TST value drawn from its generator function. In the bottom row, the same cohorts are used, but each subject contributes their mean of 7 nights. See Supplemental Figure S4 for visualization of the mixtures in panels A-C blind to cohort identity.

To assess the benefit of using more than one observation to compute an individual’s TST, we repeated the simulations where each individual’s TST value is computed from the mean of 7 simulated nights (**Figure 5D-F**). The distribution histograms are now more centered around the simulated value means, as expected from this increased number of observations per individual. When the 6-hour sub-cohort is 25% of the total, this night averaging shows that observations in the 6-hour bin are now 7x more likely to have come from the true 6-hour sub-cohort, versus the 7-hour sub-cohort (**Figure 5D**). However, at 10% prevalence of the 6-hour sub-cohort, an observation in the 6-hour bin is only about twice as likely to have come from the 6-hour as the 7-hour sub-cohort (**Figure 5E**), and at 5% prevalence (**Figure 5F**), a 6-hour observation is about equally likely to come from either sub-cohort. In summary, although repeated observations markedly improve the accuracy of a given bin assignment in this simulation, when one group is a small portion of the total cohort, observations distant from the true mean (such as “short sleep”) may be more likely to have arisen from random variations of the dominant sub-cohort (in this simulation, the 7-hour group), than from a low-prevalence group with a truly shorter mean TST value. This can also be challenging to visualize in population level histograms that contain mixtures of sub-groups, which can resemble simple Gaussians even when simulated using mixtures of two known distributions (**Supplemental Figure S4**).

Real-world TST observations allow exploration of this phenomenon more directly, again taking advantage of longitudinal data defining the “truth” for each participant. We computed the mean TST using at least 90 nights of sleep duration data per participant, which is taken as ground truth, against which the random sub-samples are compared. **Figure 6A** shows the distribution of TST values using this approach, in 1 hour bins. We drew random sub-samples of 1, 7 or 14 nights (**Figure 6**, **B-D** respectively) from participants in each of 4 “true” bins of 5 to <6 hours, 6 to <7 hours, 7 to <8 hours, and 9+ hours. The results illustrate how intra-individual variation can lead to bin mis-assignment (bins filling outside of the “truth” category), which is mitigated by increasing the duration of observations (narrower distribution of values, fewer of which land outside of the true bin).

**Figure 6.**
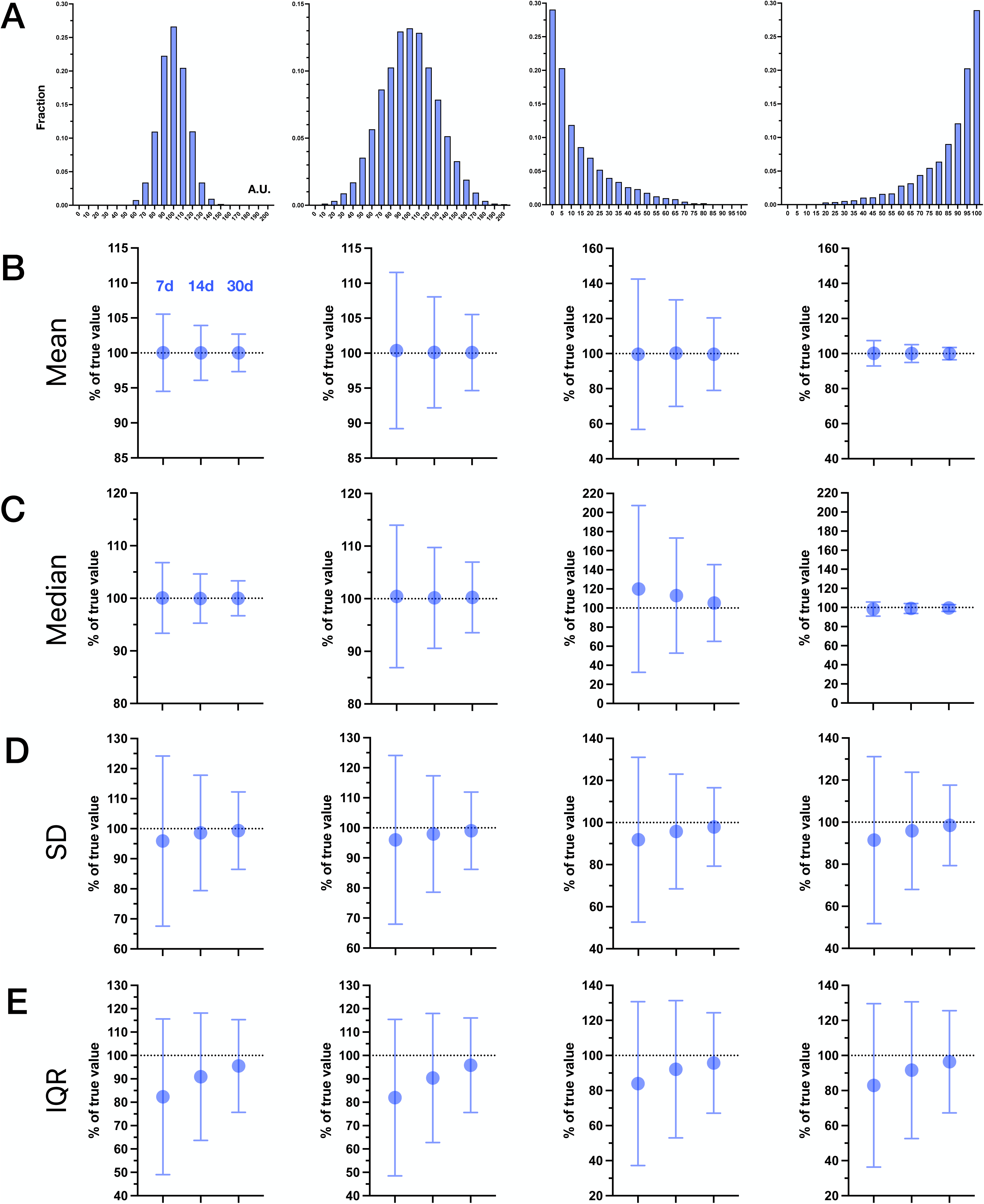
Distributions of TST values from real-world data at varying sub-sampled observation windows. A: Ground truth TST binning of the real-world cohort, using 90+ nights of TST to calculate each participant’s mean TST. Due to small samples in extreme bins, the <5 and >9 bins are not shown. In panels B-D, histogram distributions are shown for TST durations, one value per participant, calculated from sub-samples of 1 (B), 7 (C), and 14 (D) observations each. Each of the four columns correspond to participants classified in each of the 1-hour bins in panel A, to illustrate the spread of possible observations among individuals assigned ground truth bins of 5-6, 6-7, or 7-8 hours (columns) using 90+ TST values, when a random sub-sample is taken (rows). The orange dotted line is the median value. The X axes in B-D are TST values (hours).

### 4. Discussion

This study utilized a combination of simulations and real-world longitudinal data to provide an inferential framework around the limitations of short duration sampling of temporally variable data types such as sleep duration. We found that directional biases and uncertainty ranges depend on three main factors: a) the sample size of observations within each participant, b) the descriptive statistic used to capture centrality or dispersion, and c) the distribution shape of the data (Gaussian or skewed). Simulations with known distributions, as well as real-world sleep and exercise data, each demonstrated limitations of sub-samples in the form of uncertainty ranges and/or directional bias, compared within-individual to longer sampling windows used as ground truth. Statistical tests of distribution shape carry uncertainty in small sub-sample sizes, even when simulated from known simple distributions. The results have implications for interpreting existing sleep epidemiology literature based on small observation windows (1-14 days), for understanding potential differences among studies, and in planning analytics for the increasingly common setting of longitudinal cohorts with repeated measures from wearable devices.

#### 4.1 Observational studies using objective sleep metrics

It is not uncommon to use one or a few nights of PSG data to draw inferences across a range of research questions. Normative data regarding sleep metrics such as TST and sleep staging has been published in two meta-analyses, which drew mainly from studies of 1-3 nights of PSG data, although the two studies differed in whether they included^2^ or excluded the first night^1^ when more than one PSG night was available. Research studies regarding insomnia with objective short sleep used single night PSG data per assessment, such as the Penn State cohort^12^, as well as the Sleep Heart Health Study cohort^13^. Observational cohort studies with actigraphy-based sleep tracking often use 7 or 14 days. For example, the UK biobank cohort includes 7 days of actigraphy^14^, as does the Multi-Ethnic Study of Atherosclerosis (MESA^15^), and the Jackson Heart Sleep Study^16^. These cohorts have resulted in publications regarding the health associations of sleep duration and sleep variability^17–20^ (although other studies failed to show health outcome correlates with sleep, such as the HypnoLaus cohort^21^).

Key limitations when linking sleep metrics to adverse health outcomes include assessment method (e.g., PSG versus actigraphy) and the relative impact of sub-sampled observation windows both from a statistical standpoint (as in the current work) and with respect to representativeness of the sampling window (non-stationarity). The question, “how long is long enough?”, is crucial for study planning and interpretation, but the answer may be influenced by many intrinsic and extrinsic factors. For example, the night-to-night variability of any sleep metric may be a complex combination of behaviors and the presence and vulnerability to perturbations. These sources of influence on sleep would compound any inherent (biological) stochasticity^22^ that may be observed even under constant conditions. External pressures spanning work, social, and family contexts add further variability in naturalistic settings. The combination of these factors may differ among individuals, and may differ over time within individuals, contributing non-stationarity and thus questions of representativeness for any given window, especially if short duration. Ultimately, the practical consequence is that representativeness of an observation window is unclear, without having longer-term data (in which case, shorter windows would not be utilized). Longitudinal tracking overcomes some of the limitations associated with relatively short observation windows, but inter-individual variability needs to be recognized even when observations are relatively long. In the current analysis of AHMS, for example, the likelihood of passing tests of normality or log-normality, two common statistical assumptions applied to cohort data, showed heterogeneity and, in fact, failing both tests was the most common observation in the empiric data for sleep and exercise even with six months of observations (**Figure 4**).

#### 4.2 Studies of intra-individual variability

Bei, et al. summarized numerous studies that tracked sleep with actigraphy, most of which had durations of 7-14 nights^3^. Actigraphy has well known limitations in sleep-wake classification, with typically high sensitivity for sleep but low specificity^23^, such that TST is over-estimated in proportion to the amount of true wake, that is, as sleep efficiency decreases. Even if actigraphy were as accurate as PSG, our current results illustrate statistical and conceptual limitations apply to the interpretation of intra-individual variability (IIV) obtained from relatively short observation windows. In their meta-analysis, Bei, et al. reported that the SD of sleep duration was the chosen metric used to describe sleep variability in 65% of studies. Our results demonstrate that, under the Gaussian distribution assumption, the SD will under-estimate the true value at 7 days, and will vary from the true value by more than 25% (one SD) in ∼32% of cases (**Figure 1**), and that similar under-estimation occurs when analyzing empiric data from AHMS participants (**Figure 2**).

Within-individual sleep variability has been linked to a variety of adverse health outcomes, including mental health^24^, cardiovascular health^17^, metabolic health^25^, and obesity^26^. If variability is assessed with one week of sleep data, our results suggest that underestimation of SD or IQR will occur even if the underlying process is a simple normal distribution, and this underestimation may be accentuated in conditions of skewed distributions. Both the uncertainty of the estimate, and the under-estimation bias of the SD, are predicted to bias toward the null in studies attempting to link higher SD to negative health outcomes, such that associations may not reach significance, or if they do, the magnitude of association may be under-estimated. As one empiric example of this issue in the current data, the Spearman correlation coefficient between age and the SD of TST progressively decreased from a value of -0.22 using >=90 values per participant, to -0.17, -0.13, or -0.08, using 30, 7, or 3 nights per participant, respectively (data not shown).

On the other hand, uncertainty and underestimation of the true SD could also theoretically bias certain analyses toward an inflated effect, when considering attempts to control for confounders. If confounder effects are under-estimated due to uncertainty and/or under-estimation of SD, and thus incompletely accounted for in a model, this could bias toward over-estimation of the importance of sleep variability. For example, if a demographic factor such as age were associated with an outcome such as hypertension, as well as an exposure such as sleep variability, then short duration sleep tracking could result in the true relation of age to sleep variability being under-estimated, leading to incomplete control (age as a confounder), such that the relation of sleep variability to hypertension is over-estimated.

The specter of non-stationarity is likely in sleep data, given the myriad of factors known to impact sleep on any given night, and the fact that those influences themselves may vary over time. Consider the seemingly simple issue of weekend versus weekday sleep behavior differences, which could impact descriptive statistics computed from a short sampling window. A single week of TST observations divided are essentially even further sub-sampled with respect to the weekend and weekday subsets (two and five night samples, respectively). **Supplemental Figure S5** shows a simulation of how a longer weekend TST (8 hours) compared to weeknight TST (7 hours) impacts the sub-sampling approach. Specifically, a weekend change of this kind causes abrupt changes in a rolling SD computation. Even if longer duration sleep on the weekend is an isolated “event”, and other weekend nights follow the same distribution as weekdays, it takes many weeks for the cumulative SD to settle back within 5% of the true value. Influences on sleep duration that have long effect lag-times, or that themselves change slowly, could require months or longer to assess, such as seasons, changes in weight, or adoption of a new habit that takes time to master. These and other transient perturbations (e.g., illness) may be analyzed through the lens of change-point analysis.

#### 4.3 Matching statistical assumptions to phenotypes of interest

It is commonly understood that confidence in any computed statistic is proportional to sample size. However, in short duration studies of sleep, confidence bounds and error bars are more likely focusing on the population sample size, rather than on the computed sleep metrics per participant (mean, SD, etc). Explicit accounting for this latter issue is important, since it can bias hypotheses toward or away from the null, depending on the circumstances. Even the choice of descriptive statistics can be challenging, given that the within-participant true distribution cannot be confidently understood with <14 nights (**Figure 4**). One option to mitigate is to assess comparative goodness of fit metrics to choose the more favorable fit. While metrics such as the median and IQR may be chosen as a conservative approach not involving a normality assumption, it is worth noting that the IQR shows more prominent under-estimation than the SD, even for a true Gaussian distribution (**Figure 1**).

Understanding distribution shape also has implications for handling outliers. Absent a strong prior belief as to the true expected distribution, it would be difficult to interpret an apparent outlier point in a small sample, which could easily be part of a tail of a skewed distribution that was simply too sparsely sampled to properly visualize the tail. Even in settings where prior information is available at the population level (or even perfect knowledge, i.e., from a simulation), the current results show that distribution shape exhibits individual heterogeneity (see pass rates for tests of normality and log-normality in **Figure 4**).

Assigning a category label, such as “short sleep” or any sleep duration bin, carries related challenges. Simulations demonstrate how population-level questions play out, such as, given a specified sleep distribution, what is the probability of observing sleep of a particular duration? Simulations also inform the inverse conditonal question, that is, given an observed sleep duration, what is the probability it came from a short-sleeper? Answering that question requires specifying not just the distribution parameters, but their relative prevalence in the population. Neither of these is known in human observational studies. Label mixing, as seen in simulated and empiric data regarding short TST bins, could lead to under-estimation of health associations of short sleep.

Ultimately, the question of how long to track sleep depends on the goals of analysis as well as the statistical characteristics of the actual sleep measurements for each individual. Many common factors contribute to sleep patterns deviating from simplifying assumptions, such as weekends, work schedules, monthly cycles, seasons, and numerous other episodic transients (pain, mood, stress, life events, family obligations). Larger observation windows increase confidence in assessing distribution shape, and reduce the biases described herein regarding short observation windows. They also allow more complex analysis of stationarity or change-point approaches to identify periodic (e.g. weekend or monthly) or non-periodic fluctuations in the data (e.g., transient illness).

#### 4.4 Limitations

The current analysis is based on a subset of the AHMS cohort, and the extent to which the results about distribution shape and heterogeneity generalize to other populations, or using other tracking technologies, is unknown. However, the measures of variability of TST obtained in this cohort, as measured by Apple Watch first party sleep tracking, are similar to the range reported in other studies (e.g., SD of TST in the range of 1-1.5 hours). The relevance of statistical principles associated with relative-under-sampling are demonstrated via simulations, meaning, the uncertainties and biases associated with small samples can occur even under perfect measurement information assumptions. We did not address the potential for non-stationarity of sleep or exercise habits over time. Non-stationarity is likely common, and is predicted to increase measures of variability and complicate measures of centrality, if the measurement window for any individual is not considered stationary.

## Data Availability

The aggregated data that support the findings of this study can be made available on request from the corresponding author (M.B.). Request for data will be evaluated and responded to in a manner consistent with the specific language in the study protocol and informed consent form.

## Acknowledgements

The authors acknowledge the important data contributions provided by all participants in the Apple Heart & Movement Study, without whom this research would not be possible. We also thank the following individuals for their thoughtful discussion, helpful guidance, and other valuable efforts in support of this work: Simon Saffer, Tom Furman, Callum McRae, Angela Spillane, Jeff Stein, Ian Shapiro, Jen Block, Laura Rhodes. The Apple Heart & Movement Study receives funding from Apple, Inc. and the American Heart Association.

---

**Supplemental Figure S1.**
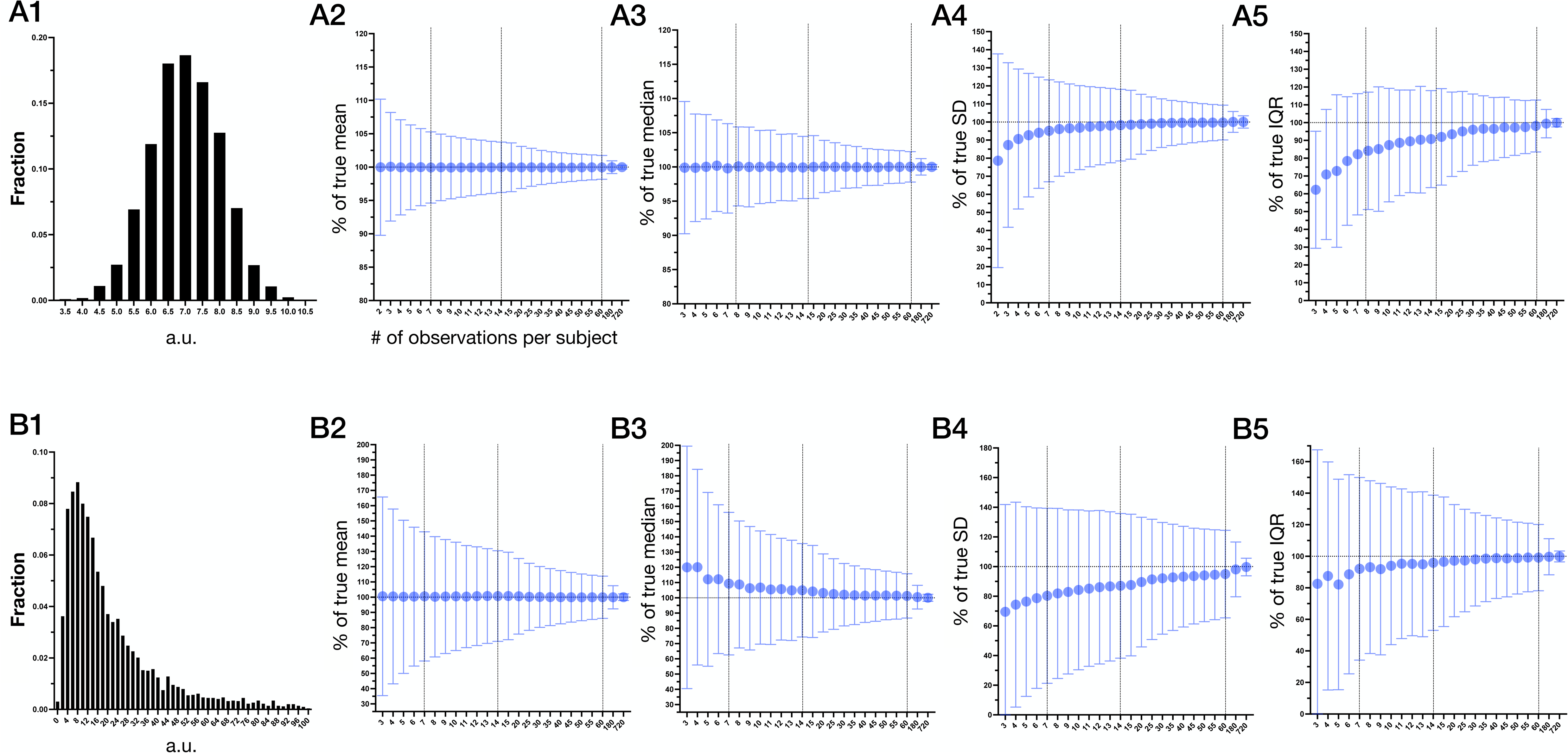
Descriptive analyses across varying sub-samples drawn from a simulated normal or log-normal distribution. Panel A1 and B1 show the distribution of 5000 random values from a normal or log-normal distribution, respectively (parameters of 7+/-1, and 2.7(0.9). In the remaining panels, the mean (circles) and standard deviation (error bars; SD) of each summary metric is given for n=5000 simulated subjects, where the X axis indicates the number of observations drawn from each simulated subject (X label in A2 applies to all panels). Note the sub-sample sizes (X axes) have non-linear increments. The vertical dotted lines are for visual convenience at 7, 14, and 60 sample windows. The summary metrics are normalized to the value obtained from n=1000 draws, taken to be the reference truth for each simulated subject.

**Supplemental Figure S2.**
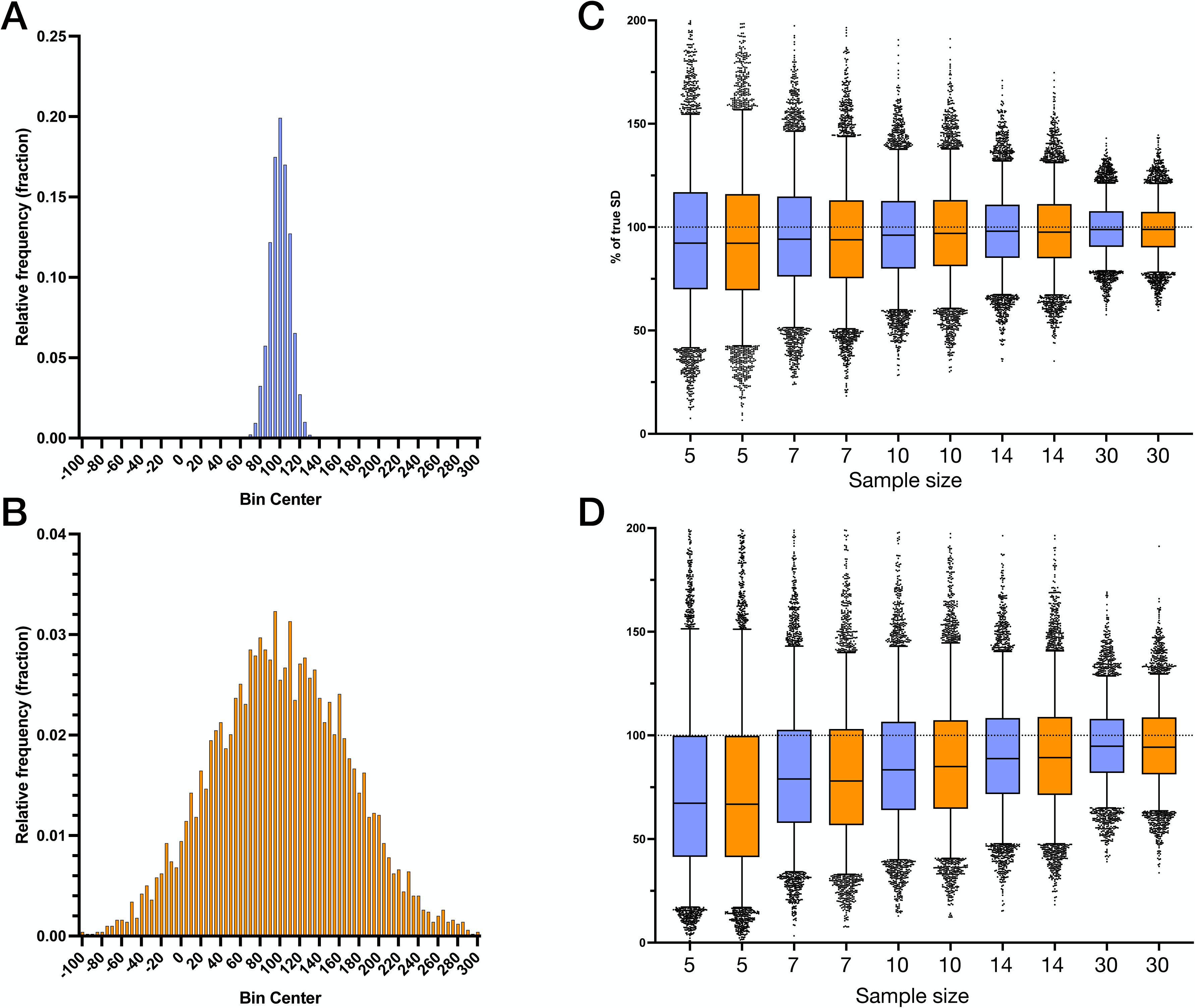
Measures of dispersion in sub-samples drawn from normal distributions with small versus large SD values. A. Histogram of 5000 values drawn from a normal distribution with mean 100 and SD 10 (a.u.). B. Histogram of 5000 values drawn from a normal distribution with mean 100 and SD 70 (a.u.). C. Distribution of SD values computed from sub-samples of the narrow (blue) versus wide (orange) distributions corresponding to panels A and B, respectively. The box plots show the median, IQR, 5-95%ile, and dots for individual points beyond the whiskers. The values are scaled as a percentage of the “true” value computed from 1000 values per simulated individual (horizontal dotted line). D. Distribution of IQR values computed from sub-samples, as in panel C.

**Supplemental Figure S3.**
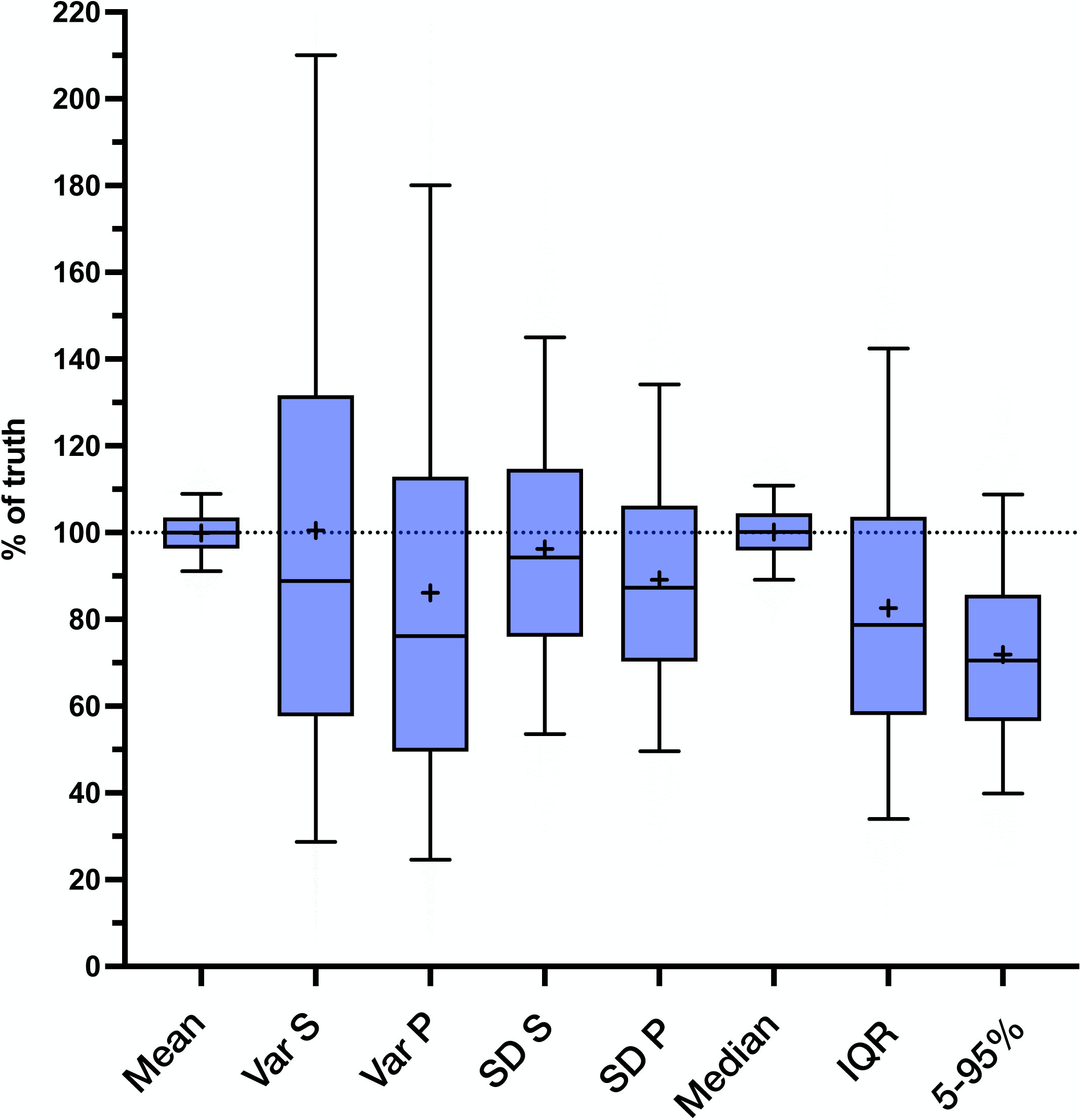
Measures of centrality and dispersion, for sub-samples of n=7 nights, drawn from a simulated Gaussian distribution. In this simulation, values are drawn from a normal distribution of 7+1 hours of sleep, for 1000 nights each of n=5000 simulated individuals. The distributions across these 5000 simulated individuals are shown as box plots for centrality (mean and median) and dispersion (variance, standard deviation, interquartile range (IQR), and 5th to 95th percentile range), in each case normalized to the true values defined by using all 1000 nights per individual. For variance (Var) and standard deviation (SD), two versions are shown for each, the population (P) and sample (S) versions of the computations (Excel command versions), which use a denominator of either n or n-1, respectively. Each box plot shows the median, IQR, and 5-95%ile (whiskers), as well as the mean (“+”), for the values obtained from 5000 simulated subjects.

**Supplemental Figure S4.**
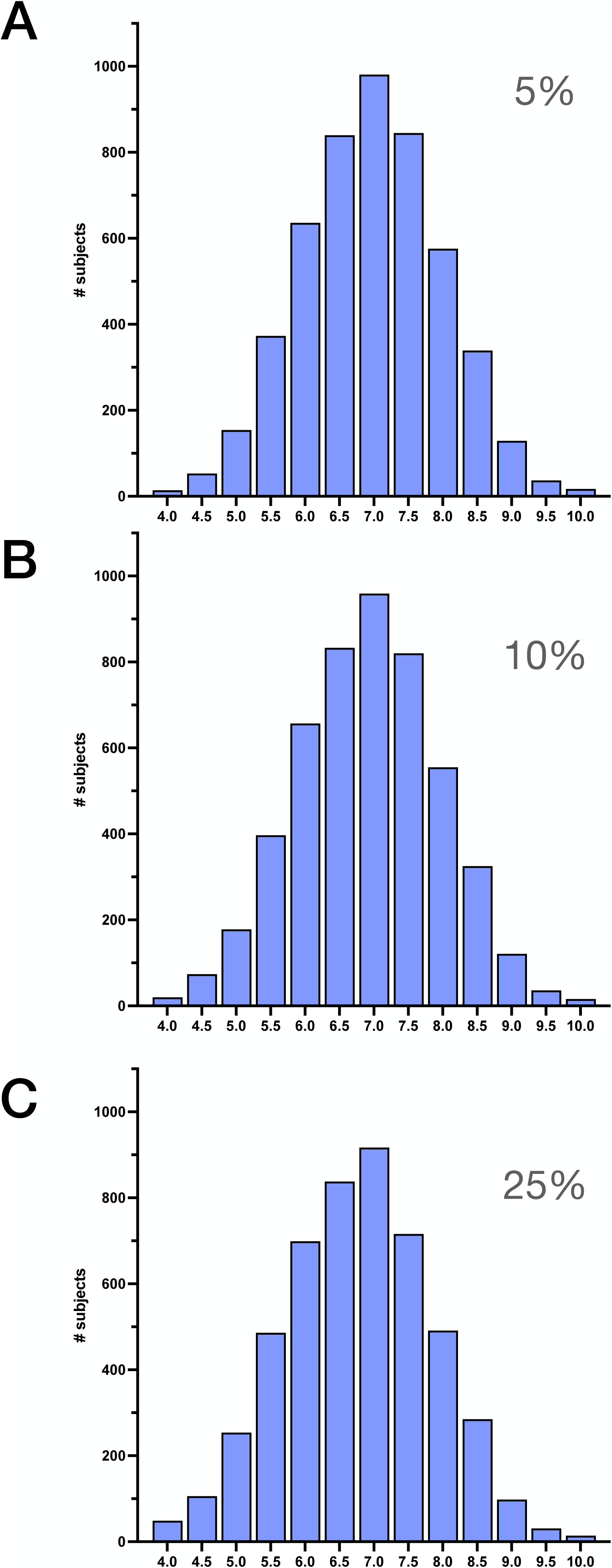
Distributions of simulated cohorts with a mixture of two normal distributions. In each panel, the binned histogram distribution is shown for simulated cohorts of n=5,000 subjects, with each subject contributing a TST value drawn from a normal distribution of either 7+/-1 hours or 6+/-1 hours, where the relative contribution of the shorter (6hr) TST is 5% (A), 10% (B), or 25% (C). (see Figure 5, for the corresponding cohorts broken out in interleaved bar graphs).

**Supplemental Figure S5.**
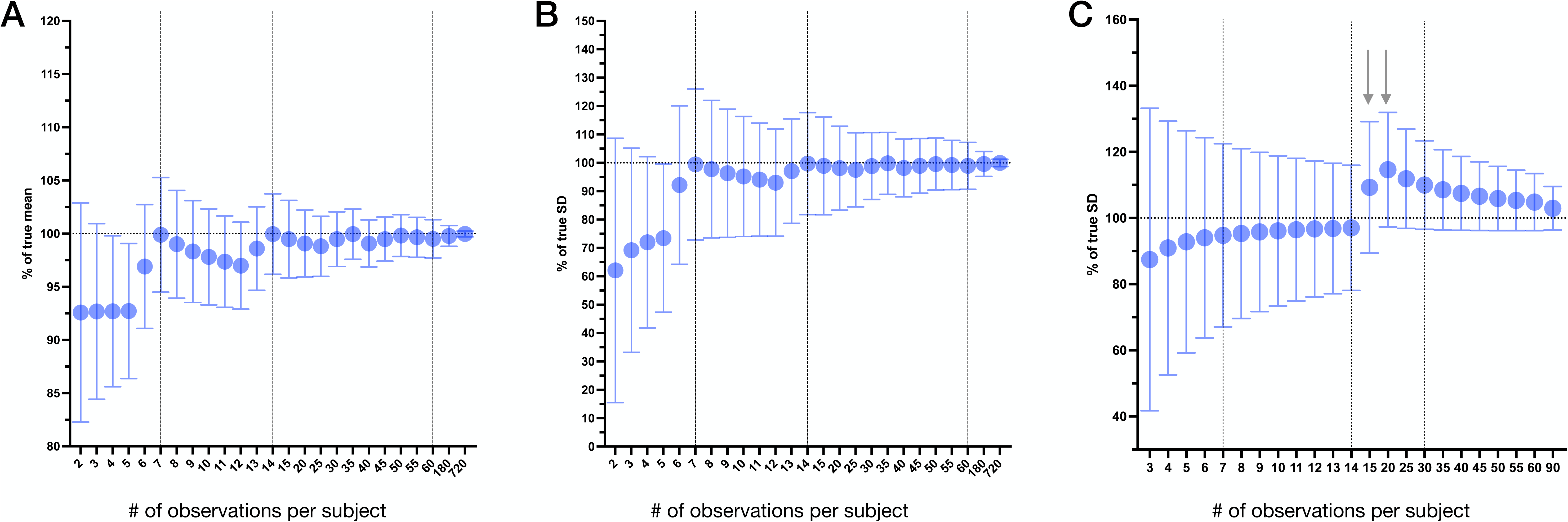
Mean and SD of sub-samples computed from a simulated cohort with longer weekend sleep. In this simulation, weekdays are drawn from a normal distribution of 7±1 hours of sleep duration, and weekends are drawn from a normal distribution of 8±1 hours. In each panel, the mean (circles) and standard deviation (error bars; SD) of each summary metric is given for n=5000 simulated subjects. The X axes in panels A and B indicate the number of sequential (not random) observations drawn from each simulated subject, beginning on a Monday such that the weekend days are 6 and 7 (when jumps are seen from the longer weekends), then dip again as more weekdays accumulate, until eventually there is convergence as more weekends are incorporated. Note the sub-sample sizes (X axes) have non-linear increments. The vertical dotted lines are for visual convenience at 7, 14, and 60 sample windows. The computed mean and SD values for each sub-sample are normalized to the value obtained from n=1000 draws, taken to be the reference truth for each simulated subject, such that the Y axes represent the % of this true value. Panel C is arranged similarly, but with 14 days of a distribution of 7±1 hours of sleep, then a single weekend drawn from a distribution of 9±1 hours of sleep (arrows), then back to 7±1 hours through day 90.

